# Validity of Computer Based Administration of Cognitive Assessments compared to Traditional Paper-based Administration

**DOI:** 10.1101/2020.05.12.20099507

**Authors:** Siao Ye, Brian Ko, Huy Phi, David M. Eagleman, Benjamin Flores, Yael Katz, Bin Huang, Reza Hosseini Ghomi

## Abstract

Traditional pen and paper based neuropsychological tests (NPT’s) for cognition assessment have several challenges limiting their use. They are time consuming, expensive, and require highly trained specialists to administer. This leads to testing being available to only a small portion of the population and often with wait times of several months. In clinical practice, we have found results tend not to be integrated effectively into assessment and plans of the ordering provider. Here we compared several tests using BrainCheck (BC), a computer-based NPT battery, to traditional paper-based NPT’s, by evaluating individual tests as well as comparing composite scores to scores on traditional screening tools. 26 volunteers took both paper-based tests and BC. We found scores of four assessments (Ravens Matrix, Digit Symbol Modulation, Stroop Color Word Test and Trails Making A&B Test) were highly correlated. The Balance Examination and Immediate/Delayed Hopkins Verbal Learning, however, were not correlated. The BC composite score was correlated to results of the Saint Louis University Mental Status (SLUMS) exam [1], the Mini-Mental State Examination (MMSE) [2], and the Montreal Cognitive Assessment (MoCA). Our results suggest BC may offer a computer-based avenue to address the gap between basic screening and formal neuropsychological testing.

## BACKGROUND

Neuropsychological tests (NPT) are standardized performance-based methods to assess cognitive functioning, and are effective in diagnosing disorders of cognition such as dementia, mild cognitive impairment, and traumatic brain injury [3]. NPT’s are typically administered in a battery, with different tests that assess a wide range of cognitive domains including memory, attention, processing speed, visuo-spatial function, and problem solving.

Traditional methods of neuropsychological assessment batteries are performed on pencil and paper. Most commonly used are screening tools with referral to formal NPT triggered in a minority of cases. Most often, NPT is considered for patients who are still in a mild stage of cognitive impairment and may have concern for an underlying neurodegenerative disorder, where NPT can offer a baseline to compare future testing to. It is also used to obtain a much more granular view than screening tools provide into a patient’s various cognitive domains which can be helpful diagnostically and for treatment planning. Screening tools common in practice include the Saint Louis University Mental Status (SLUMS) exam [1], the Mini-Mental State Examination (MMSE) [2], and the Montreal Cognitive Assessment (MoCA) [4]. However, these tests are time consuming, require verbal administration and interpretation by trained clinicians, offer only a cursory review of cognitive performance lacking sensitivity and specificity, and are not eligible for reimbursement from insurance payors. With increasing technological advancements, much work has been put into developing computerized administered NPT. Previous work on individual assessments such as the Raven Standardised Progressive Matrices test [5], trail making test A and B [6], digit symbol test [7], and stroop test [8] have demonstrated validity and reliability of computerized tests compared to the pen and paper versions. Computer-based testing has not been used in clinical practice as of yet and remains predominantly in the research space.

BrainCheck (BC) is a computerized neurocognitive testing application that is available on iPad, iPhone or desktop browser. The BC assessment interface administers neurocognitive tests, which work to maximize diagnostic accuracy, portability, and ease of operator use. Previous BC testing battery studies have been validated for diagnostic accuracy, including BC Sport for concussion [9], and BC Memory for identifying cognitive decline possibly due to dementia [10]. The primary objective of this study is to assess and validate the BC administered version of individual NPT’s compared to the traditional pen and paper versions of those tests.

## METHODS

### Comparison between BrainCheck and traditional paper-based neuropsychological assessments

BC’s validation testing involved comparing the computer-based test against the standard written form (pen and paper) of the test. Each of the participants took both BC and correlating written tests. A written test was not available for the Flanker test. The written tests that were taken by the participants include the following tests [11]:

- Ravens Matrix;
- Digit Symbol Modulation;
- Stroop Color Word Test;
- Trails Making A&B Test;
- Balance Examination; and
- Immediate/Delayed Hopkins Verbal Learning

In order to successfully validate the BrainCheck App, the acceptance criteria are:

- p-value across all tests, p<0.05
- Correlation value across all tests is > 0.60

P-value less than 0.05 indicates the BC test and the written tests are correlated. The test is also considered failed if the actual test results do not correspond to the acceptance criteria as detailed in each test section or the actual results section indicates “fail”.

### Comparison between BC test and traditional screening tools

In addition, we compared the overall composite score given by BC test [10] to results of traditional screening tools. We tested volunteers from various community centers and living facilities in Houston, Texas. Inclusion criteria included: function in at least one hand, and normal or corrected vision. Exclusion criteria included: history of stroke or other neuropsychiatric disability (eg, attention deficit hyperactivity disorder [ADHD] or epilepsy), inability to speak English or Spanish, and illiteracy, defined as inability to read the written informed consent for the purposes of this study. All participants signed consent forms approved by the Solutions Institutional Review Board. The volunteers were divided into four cohorts: a normative population and three comparison groups for the SLUMS exam, the MMSE, and the MoCA. Each comparison group completed their respective screening test and BC test.

## RESULTS

### Comparison between BrainCheck version and paper-based neuropsychological tests

A total of 27 subjects participated in comparing BC assessments versus traditional NPT, age ranged from 12-88 with a mean age of 46.9. Data were collected between 2/16/2017 and 3/11/2017. Tests were conducted without any disturbances. Table 1 shows the correlation and p-value for each set of tests, with an overall pass/fail result.

**Table 1:** Validation Test Results.

| Pen and Paper Test | BrainCheck Test | Correlation with Pen/Paper Tests and BrainCheck App (Acceptance Criteria >0.60) | p-Value (Acceptance Criteria $p < 0.05$) | Results |
| --- | --- | --- | --- | --- |
| Matrix | Matrix | 0.75 | 0.000015 | Pass |
| Digit Symbol | Digit Symbol Substitution | 0.67 | 0.0009 | Pass |
| Stroop | Stroop | 0.74 | 0.0001 | Pass |
| Trails B | Trail Making B | 0.86 | 0.0000007 | Pass |
| Balance | Coordination | 0.12 | 0.67 | Fail |
| Delayed Recall | Delayed Recognition | 0.19 | 0.32 | Fail |
| Immediate Recall | Immediate Recognition | 0.19 | 0.30 | Fail |
| Trails A | Trail making A | 0.86 | 0.00005 | Pass |

A total of 21 participants completed both the paper/pencil version of the Stroop test and the BC version of the Stroop test. BC Stroop reaction times were compared to paper Stroop scores yielding a Pearson’s correlation coefficient of -0.74. Higher paper Stroop scores were generally associated with faster BC stroop reaction times.

A total of 21 participants completed both the paper/pencil version of the Digit Symbol Substitution Test (DSST) and the BC version. A comparison of paper/pencil scores to BC reaction times yielded a strong negative correlation (r= -0.67). Higher paper DSST scores were generally associated with faster BC DSST reaction times.

21 participants completed both the paper/pencil version of the Trails A/B Test and the BC version. There was strong correlation between the two versions for both Trails A (r=0.86) and Trails B (r=0.86). Meaning, there was a very strong correlation between completion times among each version of the test.

20 individuals completed both the paper/pencil version of the Matrix Test and the BC version of the test. Number of items correct for the paper version were compared to the number correct for the BC version, yielding an r of 0.75. Meaning, there was a very strong positive correlation between the two versions of the test and number of items correct.

15 individuals completed both the paper/pencil version of the Balance test and the BC version of the test. 17 individuals completed both the paper/pencil version of the Immediate recall test and the BC version of the test. 11 individuals completed both the paper/pencil version of the Delayed recall test and the BC version of the test. Comparisons of these corresponding tests between the paper/pencil version and the BC version did not yield any significant correlation.

### Comparison between BrainCheck and traditional screening tools

In 583 subjects, we compared each BC test against the SLUMS exam, the MMSE, and the MoCA. We calculated the composite score [10] to compare with these screening tools. Although individual assessment correlations were only weak to moderate in strength, the composite score showed strong correlations (Figure 4). These results have been published on JMIR[10].

## DISCUSSION

The validation test findings demonstrate several BC assessments evaluate cognitive function with similar performance to their respective standard written (pen and paper) tests. The results comparing these two show that five out of eight tests - Matrix, Digit Symbol Substitution, Stroop, Trail Making A, and Trail Making B - met acceptance criteria while the Coordination, Immediate Recognition, and Delayed Recognition tests did not. Additionally, the value of utilizing BrainCheck as a digital tool in neuropsychological testing is supported by the validation of BrainCheck’s composite score as a test tool in cognitive impairment in older adults[10].

Five out of eight tests met the acceptance criteria of a p-value less than 0.05 and correlation above 0.60. The correlation was strongest in both Trail Making A/B tests with a correlation value of 0.86 and 0.78, respectively, with corresponding p-values <0.005. BC Coordination, Immediate Recognition, and Delayed Recognition did not show any significant correlation with their pen and paper versions. When interpreting Figure 1, this appears likely due to a ceiling effect, meaning that many individuals scored at the highest possible level on the recall tests and the scale does not allow scores above a certain level, therefore clustering individuals at the same level. This is often due to the test not being tuned well to the population and not offering enough of a challenge. We would hypothesize if the tests were more challenging, we would see a better spread of performance with correlations that may then meet our acceptance criteria. Motor coordination or balance is challenging to measure and the lack of correlation may be due to the inherent difference in the test. The traditional balance test focuses on balance with variations in stance, such as double-leg, single-leg, or heel-toe. The BC test extrapolates balance using the movement detection hardware in the iPad which mainly reflects motion in the upper body. It is possible the BC balance test is measuring a different motor performance than the traditional test. Traditional balance tests may also have relatively poor consistency. A systematic review of the Balance Test showed some studies reported reliability coefficients below clinically acceptable levels [12].

**Figure 1.**
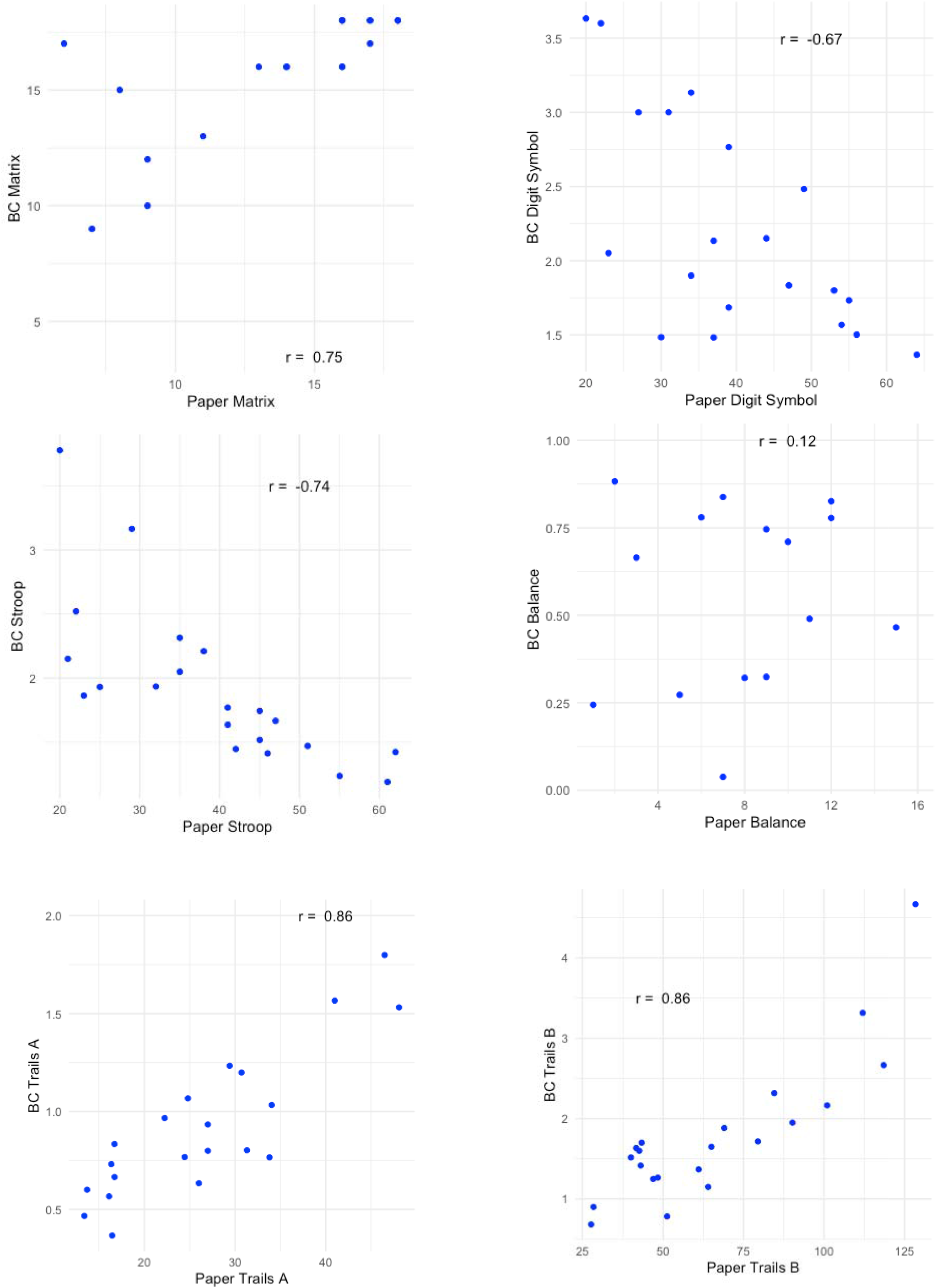
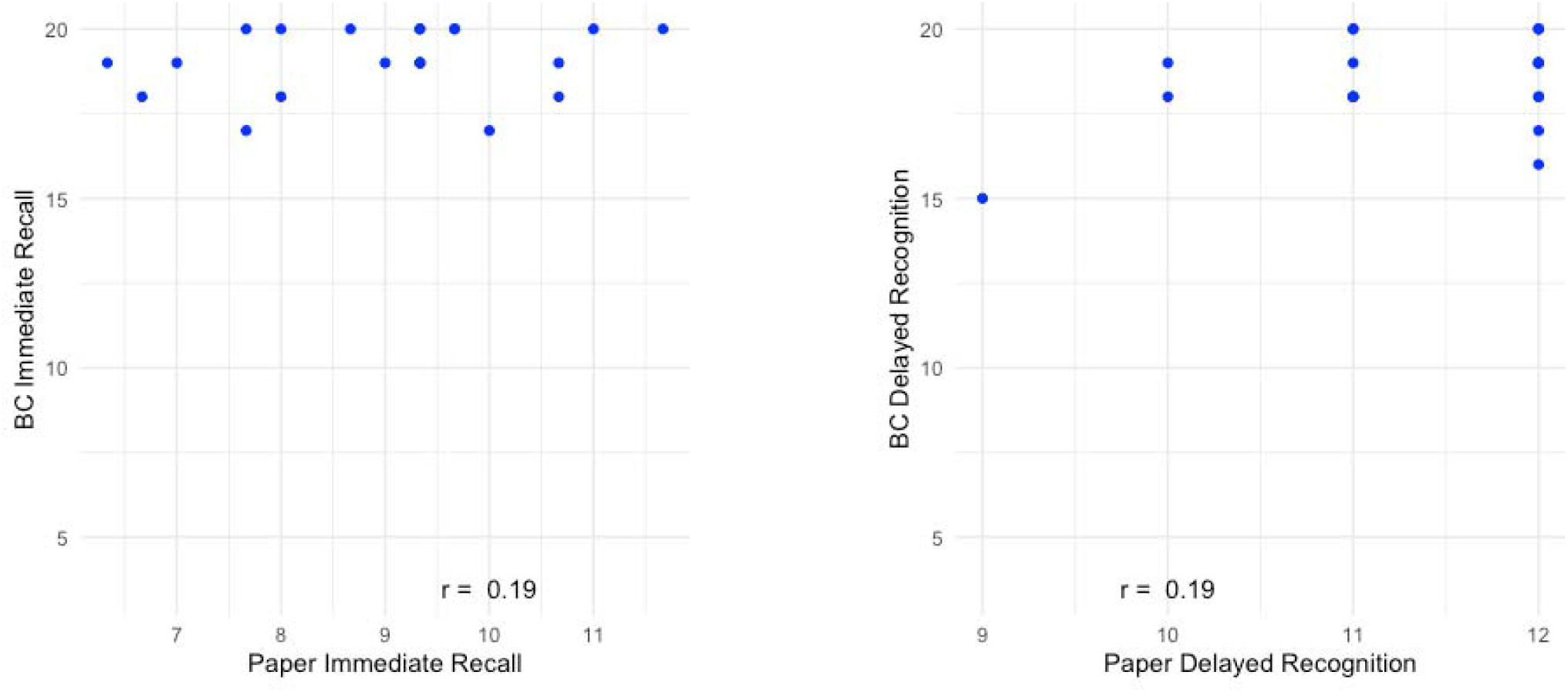
Scatter plots of paper based assessments vs. Braincheck assessments.

Convenience sampling was used to recruit participants and accordingly, our statistical power was limited by sample size. A larger cohort may have resulted in more robust results. While the large age range (12-88) may reflect the general population, the lack of stratification in age groups in addition to potential comorbidities could have resulted in inconsistencies. Thus, including stricter selection guidelines to screen for visual impairment or movement disorders that may have hindered the individual’s ability to complete the assessments could have helped provide stronger evidence. Lastly, some participants were unable to complete the entirety of BC assessments which could have contributed to limited statistical power. In the future, BC should be compared to traditional tests in larger cohorts and representative of diverse populations.

While there are numerous pen and paper tests that attempt to measure different neurocognitive domains, there is no universally accepted gold standard. Our results show that BC offers an alternative, reliable solution to augment standard written tests. The strong correlations in composite scores with other screening tools shown in Figure 2 further validates BC performance. People earned higher BC composite scores were more likely to perform well in these traditional screening tools. These findings integrated with the qualities of portability, accessibility, and low-cost of digital testing illustrate the utility of implementing computerized tests in clinical practice.

**Figure 2.**
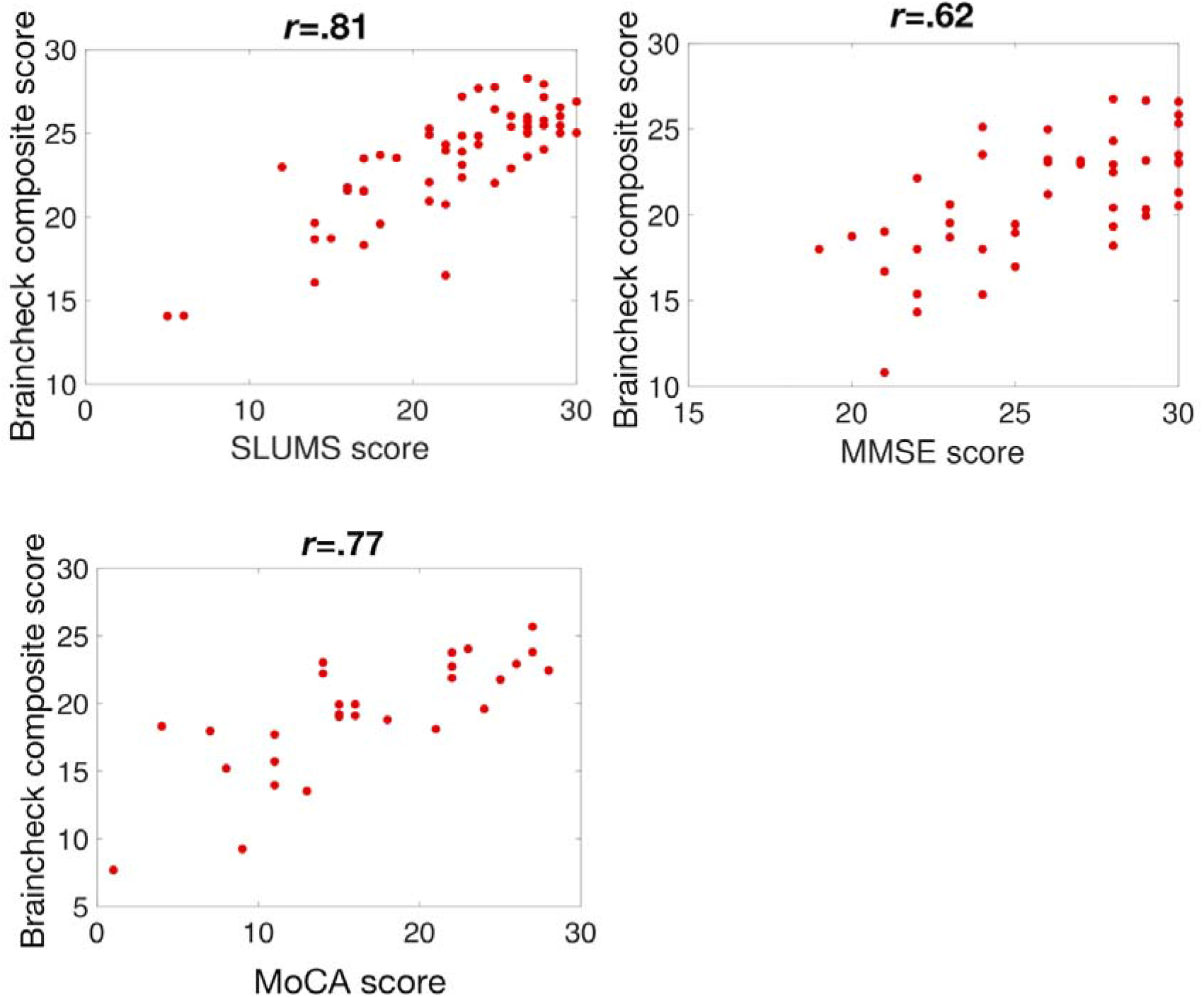
Comparison between BrainCheck composite score and the Saint Louis University Mental Status (SLUMS) exam, the Mini-Mental State Examination (MMSE), the Montreal Cognitive Assessment (MoCA).

## Data Availability

Data may be made available by contacting the corresponding author and with a data use agreement.

## Acknowledgments

Funding was provided by BrainCheck, Inc.

## Data Availability

Data may be made available by contacting the corresponding author and with a data use agreement.

